# Association of Socioeconomic Disparities and Predisposing Factors with Higher Prevalence of Hypertension related Left Ventricular Hypertrophy in Males: a Malaysian Community-Based Study

**DOI:** 10.1101/2022.04.18.22273987

**Authors:** Julia Ashazila Mat Jusoh, Thuhairah Abdul Rahman, Nafiza Mat Nasir, Norlaila Danuri, Fadhlina Abdul Majid, Fashieha Basir, Siti Norlela Ahmad Pare, Hoh Boon-Peng, Khalid Yusoff

**Affiliations:** Institute of Medical Molecular Biotechnology, Faculty of Medicine, Universiti Teknologi MARA, 47000 Sungai Buloh, Malaysia; Clinical Pathology Diagnostic Centre Research Laboratory, Faculty of Medicine, Universiti Teknologi MARA, Sungai Buloh Campus, 47000 Sungai Buloh, Selangor, Malaysia; Integrative Pharmacogenomics Institute (iPROMISE), Universiti Teknologi MARA, Cawangan Selangor, Kampus Puncak Alam 42300, Selangor, Malaysia; Primary Care Medicine Discipline, Faculty of Medicine, Universiti Teknologi MARA, 47000 Sungai Buloh, Selangor, Malaysia; Centre for Translational Research and Epidemiology (CenTRE), Faculty of Medicine, Universiti Teknologi MARA, 47000 Sungai Buloh, Selangor, Malaysia; Faculty of Medicine and Health Sciences, UCSI University, UCSI Hospital, 2, Avenue, 3, Persiaran Springhill, 71010 Port Dickson, Negeri Sembilan, Malaysia

**Keywords:** Left Ventricular Hypertrophy, population, echocardiography, hypertension

## Abstract

Left Ventricular Hypertrophy (LVH) is a risk for various cardiovascular events among those with hypertension (HT). However the prevalence of hypertension-related LVH (HT LVH+) in communities with lower socioeconomic status (SES) is not adequately reported. This study investigated the prevalence of HT LVH+ among the urban and rural males and the attributing factors. A total of 1,923 males who had echocardiographic examinations done were recruited. Their blood pressure was measured to diagnose those with or without hypertension. Left ventricular mass index was determined. Univariate analysis was performed to identify associated factors predisposing to LVH. A total of 992 males had HT, of which 264 had LVH, and were more prevalent in older age groups and Malays (p<0.001). Individuals from rural areas, with low income and low educational background were associated with higher LVH prevalence (p<0.001). Those with moderate aortic regurgitation was 3.17-fold higher in LVH. Ninety-nine normotensives had LVH, 71.7% came from rural. Total cholesterol and low density lipoprotein cholesterol levels were significantly higher in HT LVH+ from urban than the rural areas (p=0.029 and p=0.002, respectively). A quarter of the HT population in Malaysia develop LVH, majority of them were from rural, indicating that socioeconomic disparities contribute to the higher risk of HT LVH+. The rural populations may have attributed to different risk factors as opposed to those from urban, hence emphasize the need to deliver targeted strategies for prevention and management HT LVH+ by different SES.

## INTRODUCTION

Approximately 17.9 million people died from cardiovascular diseases (CVD) annually (World Health Organization; https://www.who.int/health-topics/cardiovascular-diseases/#tab=tab_1). This is more in the low and middle-income countries.^1^ In Malaysia, rural populations shows a higher prevalence of cardiovascular risk factors compared to their urban counterparts, further compounding the situation.^2^

Left ventricular hypertrophy (LVH) is an independent risk factor for cardiovascular events including heart failure, cardiac arrhythmias, strokes and cardiovascular mortality.^3,4^ Hypertension (HT), the most common cause of LVH, affects 30.8% of Malaysian male adults (aged ≥18 years old) (http://www.iku.gov.my/nhms-2019). More than one-third of HT patients developed LVH at the time of their diagnosis.^5^ Indeed, echocardiography (ECHO) study suggests that the prevalence of LVH among hypertensive males were 36.0-43.5%.^6^ Other clinical conditions that can lead to the development of LVH include valvular heart disease, heart failure, cardiomyopathy and athletic heart syndrome with physiological LVH.^7^

The prevalence of hypertensive related LVH in Southeast Asia in particular Malaysia, has not been adequately studied, except for a handful of hospital-based reports. The emergence of CVD as a leading cause of death in Malaysia runs parallel with the rapid economic growth and associated socio-demographic changes that have occurred over the past few decades. It is thus crucial to investigate the impact of urbanization and economic development on cardiovascular risk, in this case, HT related LVH. In this study, we aimed to determine the prevalence of LVH in hypertensive males among the urban and rural participants of a large community-based prospective study namely, the REsponDing to IncreaSing CardiOVascular disEase pRevalence (REDISCOVER) Study,^8^ and its attributed factors. At the time of the preparation of this manuscript, 1,923 males from REDISCOVER had an ECHO examination. Hypertensive individuals were identified from this cohort, and subsequently the attributing risk factors.

## METHODS

### Patient selection and Study Population

We included 1,923 males who had an ECHO examination through the REDISCOVER Study. ^8^ Owing to the constraints during sampling trips, the ECHO for females were limited and therefore not included in this study. Informed written consent of the participants was obtained. Demographic data was recorded. The sampling localities are shown in **Additional File 1**. Urban and rural areas were defined according to the Malaysian Population and Housing Census 2000 (https://www.dosm.gov.my/v1/). Gazetted areas with a combined population of ≥10,000 were identified as urban; whereas areas with a population of <10,000 were classified as rural.

BP was measured on a single occasion with three readings at least 1 minute apart, using a calibrated automated digital BP machine (Omron, Japan). The mean of three BP readings were taken as final reading. Height, weight, waist and hip circumference, plasma glucose and lipid levels were measured. The following criteria were applied during participant’s recruitment:

a. Age 30-80 years old.
b. HT, defined as ≥140 mmHg systolic and/or ≥90 mmHg diastolic BP, or on anti-hypertensive medication or history of HT.
c. Glucose, defined as Diabetes: Fasting Glucose (FG) ≥7.0 mmol/L (Impaired Fasting Glucose, IFG: FG 6.1-6.9 mmol/L); and Normal: FG <6.1 mmol/L.
d. Total cholesterol (TC), defined as Optimal when TC ≤5.20 mmol/L, High TC >5.20 mmol/L.
e. Triglyceride (TG), defined as Optimal when TG <1.70 mmol/L), High when TG ≥1.7 mmol/L.
f. Economic status, defined as low, middle and high according to annual incomes of ≤MYR10 000, >MYR10 000-MYR50 000, and >MYR50 000 per annum.^9^
g. Household group, defined by the Statistics Department of Malaysia (https://www.dosm.gov.my/v1/index.php?r=column/cthemeByCat&cat=120&bul_id=TU00TmRhQ1N5TUxHVWN0T2VjbXJYZz09&menu_id=amVoWU54UTl0a21NWmdhMjFMMWcyZz09) as T20 (Top 20% of median income), M40 (Middle 40% of median income) and B40 (Bottom 40% of median income).
h. Non-smoker, defined as one who has never smoked; a smoker was a current smoker. A previous smoker was one who had given up smoking.
i. BMI groups, categorized as Underweight if BMI <18.50, Normal 18.50-22.99 and overweight 23.00-27.49 and, obese if BMI≥27.50.
j. Body Surface Area (BSA) and Waist-Hip Ratio (WHR) were also obtained.
k. Cumulative number of associated factor, defined as the sum of the following factors: (i) BMI ≥23kg/m^2^; (ii) WHR >0.9; (iii) Smoking status = smoker, (iv) LDL ≥3.4 mmol/L; (v) TC >5.20 mmol/L; (vi) HDL <1.0 mmol/L; (vii) TG ≥1.7 mmol/L; and (viii) Glucose >6.

### Echocardiography (ECHO)

Doppler, two-dimensional (2D), and M-mode (2D-guided) ECHO were performed following a standardized protocol. Measurement was made using M-Mode at the Parasternal Long Axis views (PLAX), calibrated and quantitated with a computerized review station equipped with a digitizing tablet and monitor overlay, using EchoPAC (GE Healthcare).

Left ventricular ejection fraction was calculated after measuring the interventricular septal thickness at end of diastole (IVSD), left ventricular posterior wall thickness at end of diastole (LVPWD), left ventricular internal dimension at end of diastole (LVIDD) and left ventricular internal dimension during systole (LVIDS). Left ventricular mass index (LVMI) was calculated by using the formula:

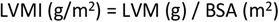

Whereby, LVM denotes left ventricular mass,

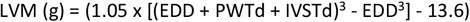

Participants were said to have LVH when in males LVMI > 125 g /m^2^.^10^

Valvular heart disease was determined based on abnormal size and/or function of the heart and a primary abnormality of a valve (i.e. presence of valve regurgitation or stenosis and thickening of cusps, leaflets, or leaflet tips, vegetations or ruptured chordae tendineae).

### Statistical analysis

Analyses were performed using SPSS (v18.0). Comparison of different risk variables between HT with-LVH (HT LVH+) and HT without-LVH (HT LVH-) were done by χ^2^-tests for categorical variables and independent samples t-test for continuous variables. Logistic regression analyses were conducted for HT and LVMI. Associations between parameters investigated and other variables were examined using Pearson’s rank correlation.

## RESULTS

The distribution of the males who underwent ECHO examination that were presented with HT and LVH is shown in **Figure 1**. Out of the 1,923 males included in this study, 992 (51.6%) had HT. Only 23.7% of the HT males achieved BP control (<140/90mmHg) while on treatment. ECHO examination showed that 264 (26.6%) of the HT participants had LVH, of which 16.3% had their BP controlled (**Figure 1**). Surprisingly, 502 (50.6%) of the study participants were not aware of having HT, 22.1% had LVH. Among the 490 participants who were aware of having HT, 26.2% were not treated and 41 (32%) developed LVH (**Figure 2**). Majority of the participants who were not aware of having HT, and those who were aware but not being treated, came from rural area with low income status (59% and 70.7%, respectively) (p<0.001)(**Table 1**). The remaining 931 samples who were normotensive (NT) during the study recruitment, 99 had LVH, 71.7% of them were from rural area. **Table 1** summarizes the demographic and conventional risk factors of the HT samples.

**Table 1:** Demographic Data and Clinical Characteristics for HT LVH+ and HT LVH-.

|  | HT LVH+<br>N=264 (26.6%) | HT LVH-<br>N=728 (73.4%) | p-value |
| --- | --- | --- | --- |
| Mean Age (years) | 61.88 ±9.66 | 57.29±10.05 | <0.001* |
| Mean Weight (kg) | 68.48±12.77 | 70.13±12.80 | 0.073 |
| Mean Height (m) | 1.61±0.06 | 1.63±0.06 | <0.001* |
| Mean BMI (kg/m<sup>2</sup>) | 26.36±4.19 | 26.29±3.99 | 0.819 |
| BMI status |  |  |  |
| Underweight+Normal | 55 (20.8) | 150 (20.6) | 0.930 |
| Overweight+Obese | 209 (79.2) | 578 (79.4) |  |
| Residential location |  |  |  |
| Urban | 71 (26.9) | 350 (48.1) | <0.001* |
| Rural | 193 (73.1) | 378 (51.9) |  |
| Mean income | 953.48±892.50 | 1552.39±1815.53 | <0.001* |
| Low income | 126 (63.6) | 236 (48.7) | <0.001* |
| High income | 72 (36.4) | 249 (51.3) |  |
| Educational background |  |  |  |
| University | 13 (5.4) | 116 (17.1) | <0.001* |
| Technic | 2 (0.8) | 13 (1.9) |  |
| Secondary school | 63 (26.0) | 265 (39.0) |  |
| Primary school | 125 (51.7) | 202 (29.7) |  |
| None | 39 (16.1) | 84 (12.4) |  |
| Occupation |  |  |  |
| Top management | 0 (0) | 12 (1.7) | <0.001* |
| Professional | 5 (1.9) | 32 (4.4) |  |
| Teacher | 12 (4.5) | 57 (7.9) |  |
| Clerk | 7 (2.7) | 30 (4.1) |  |
| Self-employed | 42 (15.9) | 104 (14.3) |  |
| Farmers | 18 (6.8) | 17 (2.3) |  |
| Art, craft and design | 1 (0.4) | 8 (1.1) |  |
| Mechanic | 0 (0) | 9 (1.2) |  |
| Agriculture | 100 (37.9) | 224 (30.9) |  |
| Security Guard | 9 (3.4) | 34 (4.7) |  |
| Retire | 38 (14.4) | 149 (20.6) |  |
| None | 32 (12.1) | 49 (6.8) |  |
| Household group |  |  |  |
| T20 | 0(0) | 5(1.2) | 0.552 |
| M40 | 10 (5.5) | 18(4.3) |  |
| B40 | 188(94.5) | 400(94.5) |  |
| Smoking |  |  |  |
| Current | 58 (22.4) | 159 (22.5) | 0.998 |
| Previous | 67 (25.9) | 184 (26.0) |  |
| Never | 134 (51.7) | 364 (51.6) |  |
| Race |  |  |  |
| Malay | 211 (79.9) | 591 (81.2) | <0.001* |
| Chinese | 7 (2.7) | 49 (6.7) |  |
| Indian | 0 (0) | 15 (2.1) |  |
| Others | 43 (16.3) | 73 (10.0) |  |
| Bumiputera | 3 (1.1) | 0 (0) |  |
| Mean Systolic BP (mm Hg) | 163.35 ±23.37 | 152.64 ±18.45 | <0.001* |
| Mean Diastolic BP (mm Hg) | 89.49 ±13.19 | 88.06 ±11.60 | 0.100 |
| TC (HYPERCHOL) |  |  |  |
| Optimal | 67 (29.5) | 163(26.7) | 0.465 |
| High | 160 (70.5) | 448(73.3) |  |
| HDL (LOW HDL) |  |  |  |
| Optimal | 152 (60.6) | 473 (68.9) | 0.017* |
| Low | 99 (39.4) | 232 (31.1) |  |
| LDL (HIGH LDL) |  |  |  |
| Optimal | 82 (56.6) | 200 (32.7) | 0.401 |
| High | 145 (43.4) | 411 (67.3) |  |
| TG (HYPER TG) |  |  |  |
| Optimal | 100 (44.1) | 255 (41.7) | 0.599 |
| High | 127 (55.9) | 356 (58.3) |  |
| Glucose |  |  |  |
| Normal | 177 (78.7) | 464 (78.6) | 0.923 |
| IFG+Diabetes | 48 (21.3) | 122 (21.4) |  |
| WHR |  |  |  |
| Normal | 63 (23.9) | 210(28.9) | 0.127 |
| Increased | 201 (76.1) | 517 (71.1) |  |
| WC |  |  |  |
| Normal | 103 (39) | 301 (41.4) | 0.511 |
| Increased | 161 (61) | 426 (58.6) |  |
| Metabolic Syndrome |  |  |  |
| Normal | 136 (51.5) | 379 (52.1) | 0.879 |
| Syndrome | 128 (48.5) | 349 (47.9) |  |

Abbreviations: BMI, body mass index; TC, total cholesterol; HDL, high-density lipoprotein; LDL, Low-density lipoprotein; TG, triglycerides; IFG, impaired fasting glucose; WHR, waist hip ratio; WC, waist circumference; T20, Top 20% of median income; M40, Middle 40% of median income; B40, Bottom 40% of median income; Case, hypertension with LVH (HT LVH+); control, hypertension without LVH (HT LVH-). \* significantly different at $p < 0.05$

**Figure 1:**
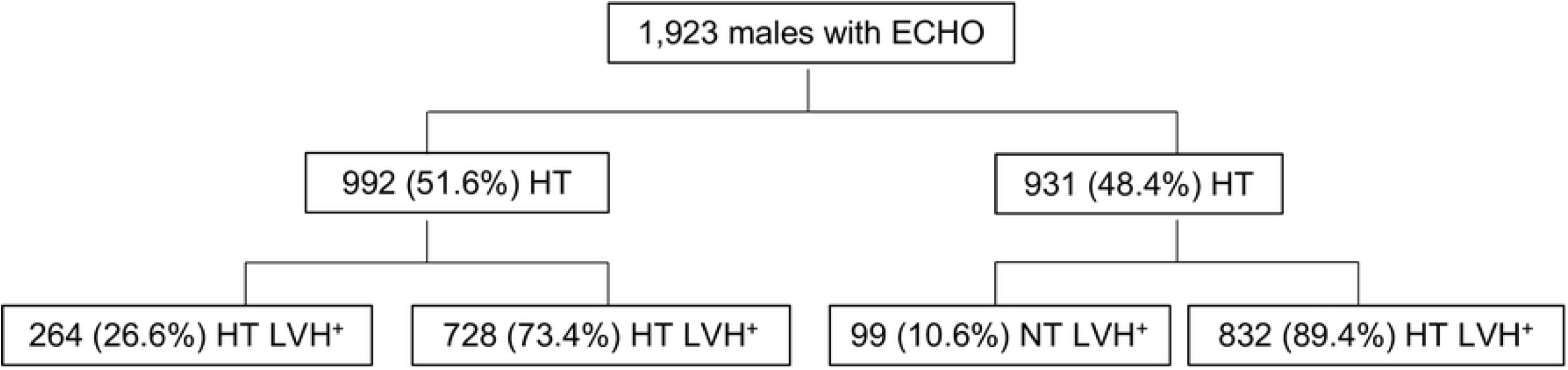
Prevalence, awareness, treatment, and control of hypertension and hypertension related LVH in the Malaysian males’ population.

**Figure 2:**
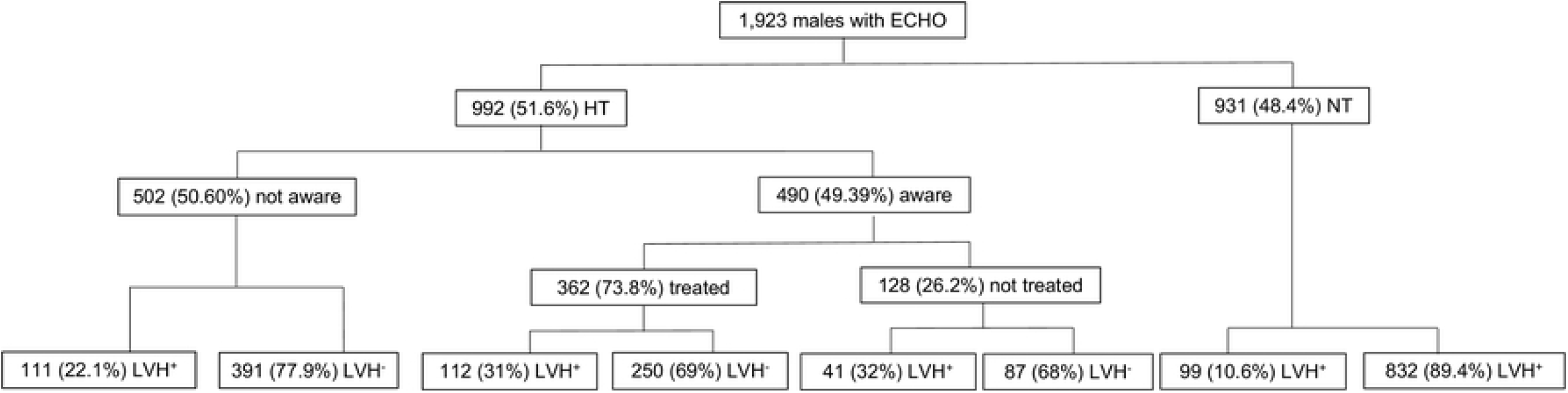
Age Distribution of Participants with HT LVH+ and HT LVH-. The peak prevalence of HT LVH+ was seen between 60-69 years old and HT LVH-was seen between 50-59 years old.

### Associated Risk Factors for HT LVH+

A weak but significantly positive correlation was found between LVMI with age (r=0.217, p<0.001) (**Additional File 2**). The HT LVH+ were on average older than the HT LVH-. Majority of them (40.7%) aged between 60-69 years old, as opposed to the HT LVH- (36.0% from age group 50-59 years old) (**Table 1**; **Additional File 3**).

Previous study reported that LVH was associated with severity of calcification of aortic valve, aortic stenosis and aortic regurgitation independent of HT, ^11,14^ therefore we asked if the same observation is found in our study population. Among the HT LVH+, 36 (32.6%) presented with calcified aortic valve and seven had aortic valve sclerosis, majority of them fell at the age group between 60 – 69 years old (**Additional File 4**).^12^ One presented with severe aortic valve stenosis and 10 (3.8%) had moderate aortic valve regurgitation, significantly higher (p<0.001) than other groups (**Additional File 6**), implying that individuals with aortic regurgitation had a higher risk in developing HT LVH+. We also found that the Malays had a higher prevalence of HT LVH+ as opposed to other ethnic groups (p<0.001).

Mean SBP was significantly higher in HT LVH+ compared to HT LVH- (p<0.001). LVMI was positively correlated with SBP (**Supplementary File 6**). We found that more HT LVH+ individuals were on treatment as opposed to HT LVH- (p<0.001). When compared to the HT LVH-, the likelihood of HT LVH+ was 1.25-fold, 1.63-fold, 1.49-fold, 4.55-fold and 5.33-fold increase in group with SBP 161-170, 171-180, 181-190, 190-200 and >200 mmHg, respectively (**Figure 3**). We found neither significant attribution between obesity and HT, nor LVMI and BMI in the HT population; however, LVMI was positively correlated with WHR (**Supplementary File 7**). HT LVH+ had lower HDL level than HT LVH- (p=0.033), and LVMI was weakly negatively correlated with the HDL level (**Supplementary File 8**). On the other hand, HT LVH+ was not associated with smoking status and the duration of smoking (≥3 years) (p=0.402).

**Figure 3:**
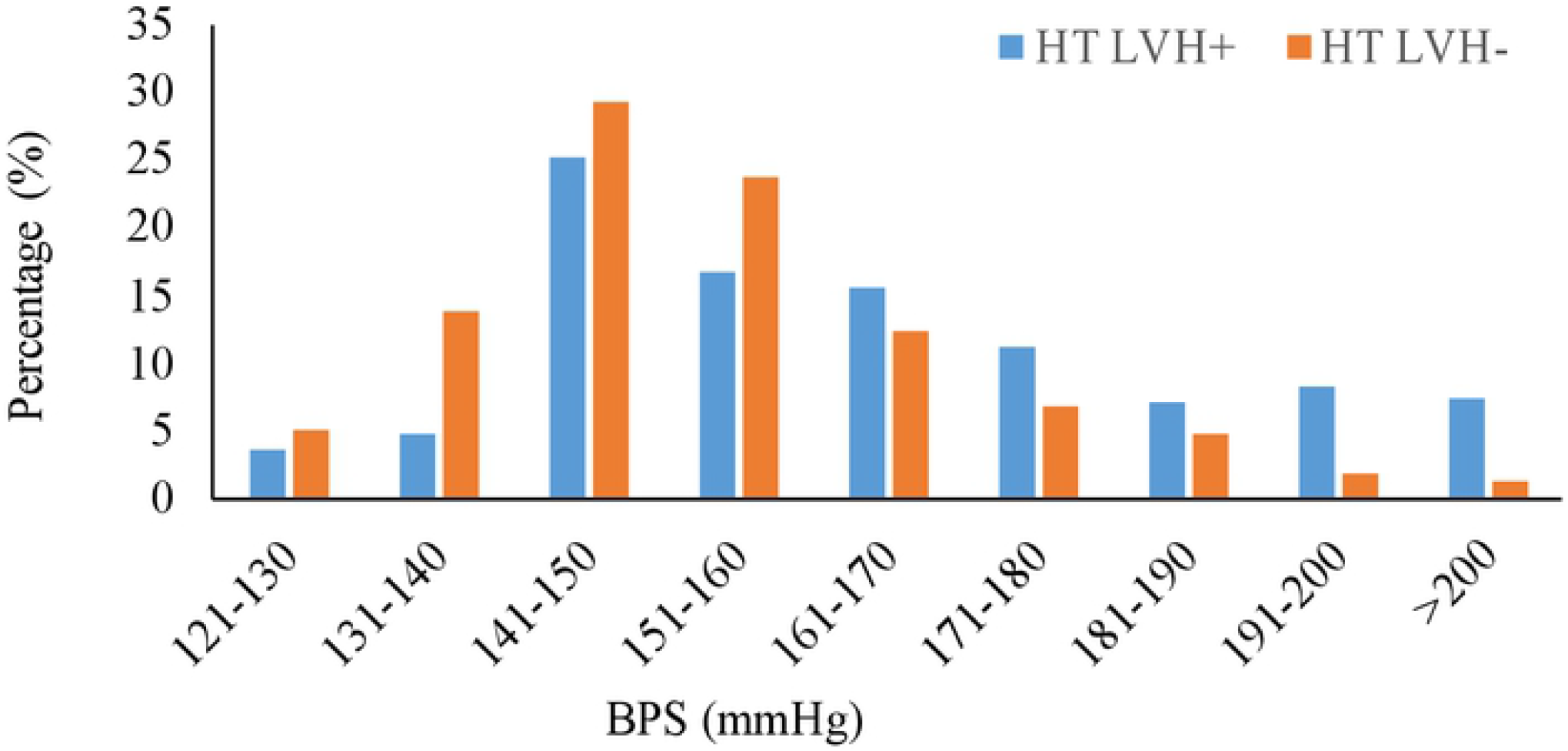
Systolic Blood Pressure (BPS) Distribution in HT LVH+ and HT LVH-. The peak percentage of HT LVH+ and HT LVH-was seen between 141-150 mmHg. Starting from BPS 161 mmHg, percentage of HT LVH+ was higher than those in HT LVH-.

### Association of Socioeconomic Discrepancies and HT LVH+

Significant lower income status was observed in HT LVH+ (p<0.001). However, when the income status was classified into T20, M40, and B40, no significant association observed between these categories and HT LVH+ (p=0.552). Individuals from rural areas had significant higher prevalence of HT LVH+ (p<0.001), and lower treatment rate (p<0.001) than the urban areas. We also revealed that the HT LVH+ in urban were older than the rural (p=0.020) (**Table 2**).

**Table 2:** Demographic Data and Clinical Characteristics among HT LVH+ Between Urban and Rural Participants.

|  | Urban<br>N=71 (26.9) | Rural<br>N=193 (73.1) | p-value |
| --- | --- | --- | --- |
| Mean Age (years) | 64.17 ±8.96 | 61.04±10.05 | 0.020* |
| Mean Weight (kg) | 70.51±11.67 | 67.73±13.10 | 0.371 |
| Mean Height (m) | 1.62±0.06 | 1.61±0.58 | 0.232 |
| Mean BMI (kg/m<sup>2</sup>) | 26.92±3.657 | 26.14±4.36 | 0.255 |
| BMI status |  |  |  |
| Underweight+Normal | 8 (11.3) | 47 (24.4) | 0.025* |
| Overweight+Obese | 63 (88.7) | 146 (75.6) |  |
| Educational background |  |  |  |
| University | 7 (12.1) | 6 (3.3) | <0.001* |
| Technic | 1 (1.7) | 1 (0.5) |  |
| Secondary school | 24 (41.4) | 39 (21.2) |  |
| Primary school | 24 (41.4) | 101 (54.9) |  |
| None | 2 (3.4) | 37 (20.1) |  |
| Occupation |  |  |  |
| Professional | 3 (4.2) | 2 (1.0) | <0.001* |
| Teacher | 6 (8.5) | 6 (3.1) |  |
| Clerk | 3 (4.2) | 4 (2.1) |  |
| Self-employed | 10 (14.1) | 32 (16.6) |  |
| Farmers | 0 (0) | 18 (9.3) |  |
| Art, craft and design | 1 (1.4) | 0 (0) |  |
| Agriculture | 10 (14.1) | 90 (46.6) |  |
| Security Guard | 6 (8.5) | 3 (1.6) |  |
| Retire | 23 (32.4) | 15 (7.8) |  |
| None | 9 (12.7) | 23 (11.9) |  |
| Mean income | 1322.06±1201.40 | 877.10±797.70 | 0.002* |
| Smoking |  |  |  |
| Current | 10 (15.2) | 48 (24.9) | 0.256 |
| Previous | 18 (27.3) | 49 (25.6) |  |
| Never | 38 (57.6) | 96 (49.7) |  |
| Mean Systolic BP (mm Hg) | 159.23 ±25.74 | 164.86 ±22.31 | 0.231 |
| Mean Diastolic BP (mm Hg) | 87.95 ±15.08 | 90.06 ±12.42 | 0.074 |
| TC (HYPERCHOL) |  |  |  |
| Optimal | 10 (17.5) | 59(33.3) | 0.0293* |
| High | 47 (82.5) | 118(66.7) |  |
| HDL (LOW HDL) |  |  |  |
| Optimal | 36 (53.7) | 118 (61.8) | 0.2514 |
| Low | 31 (46.3) | 73 (38.2) |  |
| LDL (HIGH LDL) |  |  |  |
| Optimal | 11 (19.3) | 75 (42.4) | 0.002* |
| High | 46 (80.7) | 102 (57.6) |  |
| TG (HYPER TG) |  |  |  |
| Optimal | 25 (43.9) | 75 (42.4) | 0.879 |
| High | 32 (56.1) | 102 (57.6) |  |
| Mean Glucose (mmol/L) | 6.54±3.07 | 5.94±2.64 | 0.030* |
| Glucose |  |  |  |
| Normal | 45 (65.2) | 136 (72.7) | 0.241 |
| IFG+Diabetes | 24 (34.8) | 51 (27.3) |  |
| WHR |  |  |  |
| Normal | 21 (29.6) | 42 (21.8) | 0.196 |
| Increased | 50 (70.4) | 151 (78.2) |  |
| WC |  |  |  |
| Normal | 26 (36.6) | 77 (39.9) | 0.671 |
| Increased | 45 (63.4) | 116 (60.1) |  |

Abbreviations: BMI, body mass index; TC, total cholesterol; HDL, high-density lipoprotein; LDL, Low-density lipoprotein; TG, triglycerides; IFG, impaired fasting glucose; WHR, waist hip ratio; WC, waist circumference. Case, hypertension with LVH (HT LVH+); control, hypertension without LVH (HT LVH-). \* significantly different at p < 0.05

HT LVH+ in urban area had higher glucose, LDL and TC levels (p=0.030, p=0.001 and p=0.029, respectively), and were more obese and overweight compared to those from rural areas (p=0.025). These results are consistent with the notion that urban sedentary lifestyle contributes increased risk of the development of hypertension related LVH.

Only 1.21% of the HT LVH+ had no associated factors identified (**Figure 4**). Majority of the HT males in this study had >5 cardiovascular risk factors. The percentages of HT LVH+ attributing with >4 associated factors was 1.03-fold (23.48% vs 22.81%) or 1.06-fold (50.61% vs 47.73%) higher than the HT LVH-. Our analysis revealed that the number of the associated factors identified was not correlated with the degree of LVMI (p=0.770).

**Figure 4:**
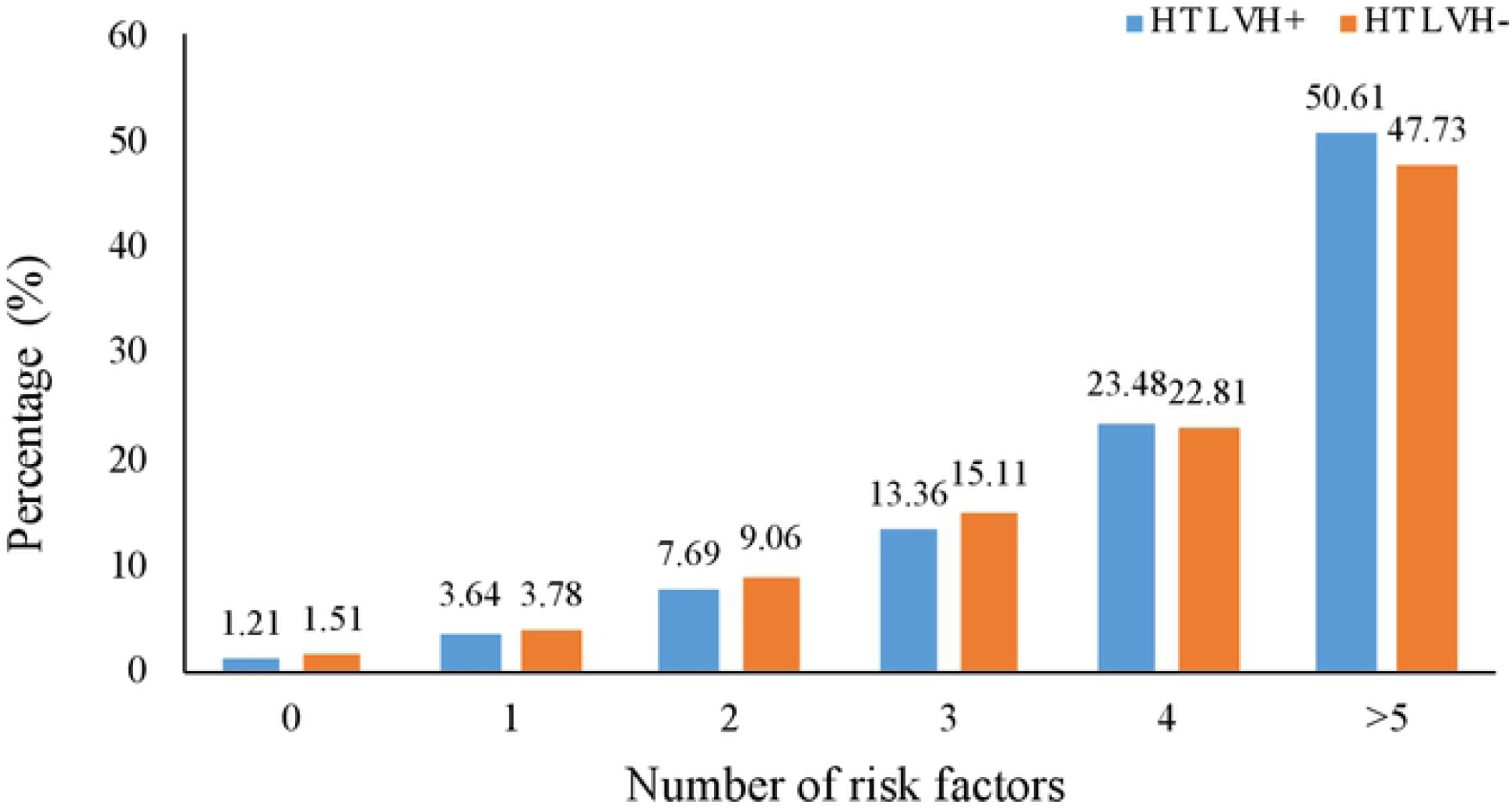
Prevalence of cumulative number of cardiovascular risk factors in HT LVH+ and HT LVH-. The proportion of HT LVH+ participants that simultaneously had four and more cardiovascular risk factors was higher than that HT LVH-participants.

Collectively the finding suggests that increased number of the cardiovascular risk factors increased the LVH risk but not the degree of LVMI.

## DISCUSSION

This study documented a high prevalence of HT (51.6%) among the male cohort aged 30 years and above, much higher than those reported from the National Health and Morbidity Survey 2019 (∼30%) (NHMS 2019; http://www.iku.gov.my/nhms-2019). The discrepancy may be due to the difference in sampling localities, and bias in ethnicities of the samples. In addition, we have also identified the associated risk factors that may contribute to the increased risk of developing LVH among the hypertensive males in Malaysia, which include age, calcification of aortic valve, SBP, WHR and low HDL level. More importantly, we showed that lower SES is more prone to adverse health outcomes, as evident by higher prevalence of HT and LVH among the communities from rural areas with lower monthly household income.

What is more alarming from the finding is that, of those with HT, half of the population were unaware that they have HT, highlighting the need to continue addressing this issue with more effective means of increasing public awareness on the significance of HT in developing CVDs and controlling BP. We also found that majority of the HT individuals, despite their compliance to the anti-hypertensives prescribed, were not successful in controlling their BP, plausibly attributed to other factors that may affect effectiveness of treatment modalities, for example, individual genetic make-up.^13^

Higher prevalence of HT LVH+ in rural areas and lower income status are possibly attributed to lack of awareness and poorer BP control in rural populations may have led to lack of knowledge on HT management, and disparity in healthcare services thence less health-seeking behaviour.^8^ We also demonstrated that NT LVH+ was more common in rural area, possibly due to the exposure of strenuous physical activity that leads to physiological response (given that farmer is the main occupation in the rural areas). Although HT is a major risk factor for LVH, the influence of physical activity should be considered. Collectively, this finding underlines the need to deliver awareness campaign targeting populations in the rural area with low-income status and low education level. In contrast, poor eating habit and sedentary lifestyle may promote increased risk of HT LVH+ in the urban populations, as indicated by higher level of LDL, TC, glucose and obesity rate. Lifestyle modification may reduce the risk of HT LVH+ development in urban participants.

HT LVH+ had lower monthly income status as opposed to HT LVH- (p=0.001); but not between household group in HT LVH+ and HT LVH- (p=0.552). However when both normotensive and hypertensive population were combined, LVH was commonly observed in the B40 group (p=0.001), suggesting that lower SES was associated with increased LVH risk regardless of BP. Low-income populations are more likely to face greater barriers to accessing medical care; yet less likely to have insurance and more likely to be employed by organization that do not offer health benefit. Policy initiative that supplements income to reduce cardiovascular morbidity and mortality is in crucial need.

The increased risk of HT LVH+ by 3-folds among the patients with aortic valve regurgitation is consistent with previous report.^15^ Furthermore, individuals with aortic regurgitation in our study also presented with aortic valve calcification, which is common among the aging population.^11^ Screening for valvular defects among HT males should be considered to reduce the risk of LVH among those found to have aortic regurgitation.

Older HT LVH+ in this study exhibited a more severe increase in LVMI, higher relative wall thickness and extra-cardiac organ damage compared with young and middle-aged sub-groups.^16,17^ As a person age, the heart undergoes subtle physiologic changes, even in the absence of disease that may contribute towards LVMI. In addition, 32.5% HT LVH+ in this study presented with calcified aortic valve – commonly observed among the elderly population and is associated with LVH.^18^

Obesity increases risk of developing LVH.^19^ It represents a state of excess adipose tissue mass. However, BMI – a common measurement for obesity – does not take into consideration regional distribution of adiposity that correlates better with cardiovascular risk. Hence waist to hip ratio (WHR) has been suggested as an alternative thus more effective predictor for cardiovascular risk compared to BMI.^20^ We found no significant correlation between BMI and LVMI in the HT population; but observed a strong correlation between WHR and LVMI, further supporting WHR as a better predictor for central obesity, as lifestyle intervention is likely to reduced risk of CVD.

Insulin has been associated with LVH owing to its trophic effects on myocytes in the heart. ^21^ Surprisingly however, we observed no association between blood glucose level and HT LVH+, consistent with the Japanese population.^22^ In the Framingham cohort study, diabetes was associated with higher LV mass in women but not men.^23^ Other factors such as obesity or elevated levels of lipid profiles that often coexist with diabetes, could possibly confound the contradicting findings between this study and those reported earlier. The HyperGEN study suggested that diabetes magnifies over time the alterations of the cardiovascular system induced by increased afterload of systemic HT.^24^ However in this study, the duration of HT was not assessed. Further studies are warranted to conclude the effects of diabetes in HT LVH+ in the context of our population.

Consistent with previous studies ^25, 26^, our study revealed lower levels of HDL-c in HT LVH+ and was inversely correlated with LV mass; but surprisingly not associated with TG level. Since obesity and serum TG were associated with increases in LV wall thickness, LV mass, and the prevalence of echocardiographic LVH independent of BP,^27, 28^ it is therefore conceivable to postulate that low HDL-c, but not TG, is independently associated with HT LVH+ among Malaysian male population.

Current smokers as well as higher levels of cumulative cigarette exposure from past smoking, were both associated with higher LV mass and LVH in the elderly. ^29,30^ Intriguingly, we found no association between the smoking status and HT LVH+ in this study. The duration and the number of cigarettes, however, are not documented in this study, therefore not able to assess its impact. Earlier study postulated that smoking contributes to insulin resistance, a possible risk factor for the development of LVH.^31^ However the exact mechanisms by which smoking contributes to alterations in cardiac structure and function is inconclusive. The lack of association between blood glucose and HT LVH+ in this study may partly explain the reason of the lack of association between smoking status and HT LVH+.

Finally, HT individuals with cumulative of four associated factors and above are more likely to develop LVH. Cardiovascular risk factors associated with HT LVH+ are closely attributed. For example, reduction of body weight may affect lipid and glucose level. Therefore, intervention with any of these risk factors may reduce likelihood of LVH development. To mitigate LVH development, it is important to identify hypertensive individuals with multiple risk factors. Recognizing and addressing these risk factors are necessary for the primary prevention and early detection of LVH in HT population.

## CONCLUSION

This study highlights three key messages: (i) a quarter of the HT population in Malaysia develop LVH, of which the highest prevalence is among the elderly; (ii) majority of the HT LVH+ were from rural areas, possibly contributed by the lower-level education and income observed in these areas, therefore supporting the notion that lower SES contributes to the higher risk of HT LVH+; (iii) the risk factors that contribute to the HT LVH+ in the urbans are different from the rural areas, hence emphasize the need to deliver strategic cardiovascular awareness campaign to targeted populations. The findings of this study provide a clearer framework for prevention and management HT LVH+ in the developing countries, particularly in the rural areas. It also provides avenues for future research to verify some of the postulations generated from this study.

## Data Availability

All relevant data are within the manuscript and its Supporting Information files.

## ACKNOWLEDGEMENTS

The authors thank the Public Health Research Institute, McMaster University, Canada as part of the REDISCOVER Study contributes to the global PURE Study. The authors also would like to acknowledge all highly dedicated research assistances and staffs from Faculty of Medicine, Universiti Teknologi MARA particularly Centre for Translation Research and Epidemiology (CenTRE), Clinical Research Laboratory (CRL), Non-Invasive Cardiac Laboratory (NICL), Institute Medical and Molecular Biotechnology (IMMB), respiratory unit and nurses involved for all their support and assistance.

## Ethical Approval

This study was approved by Universiti Teknologi MARA Research Ethics Committee [REC/UITM/2007(10)].

## Author Contributions

HBP and KY conceptualised the study. JAMJ, ND, FAM, RR, FB and SNAP conducted the experiments and were involved in data analysis and interpretation, with guidance from HBP and KY. JAMJ, HBP and KY contributed to preparation of the manuscript. All authors read and approved the final version.

**Patient consent** Obtained

**Provenance and peer review** Not commissioned; externally peer reviewed

**Data sharing statement** No additional data are available.

**Patient and Public Involvement** Patients or the public WERE NOT involved in the design, or conduct, or reporting, or dissemination plans of our research

